# Effect of a feedback loop on clinical prediction model performance

**DOI:** 10.64898/2026.11.23.26368745

**Authors:** Samantha Pacynko, Matthew Sperrin, David A Jenkins

## Abstract

Clinical prediction models (CPMs) that guide treatment-initiation, and therefore affect patient outcomes, may create feedback loops when updated using data from patients whose care they influenced. Treatment can alter predictor-outcome relationships and outcome prevalence, potentially reducing model performance. This simulation study aims to quantify this effect using setting parameters based on cardiovascular disease prediction and intervention and assess if including a treatment variable in the model mitigates any performance degradation. In the simulation, models were developed to predict the probability of a binary adverse outcome, guide treatment allocation, and then updated using data containing patients in whom the model was applied. Scenarios varied treatment inclusion as a predictor in the model and the presence of unmeasured confounding. Performance was assessed with calibration-in-the-large, calibration slope, c-statistic, and correct risk assignment relative to treatment threshold. Without unmeasured confounding, excluding treatment as a predictor resulted in miscalibration, but including treatment solves this problem completely. With unmeasured confounding, performance deteriorated even when treatment was included. These results agree with previous findings and illustrate the likelihood that models that guide interventions may experience performance deterioration due to the difficulty of recording treatment and avoiding unmeasured confounders. Causal inference methods combined with risk prediction may help address this problem. Further research is needed on monitoring techniques to allow detection of this issue post-model implementation.

## Introduction

Clinical prediction models (CPMs) are algorithms that aim to predict health outcomes based on individual patient characteristics in order to inform decisions about patient care, such as whether to start treatment or do additional screening. [1] A well-known example is QRISK3, which estimates the 10 year risk of cardiovascular disease (CVD) and is recommended for use in England and Wales. [2,3]

When models affect clinical decision-making, a feedback loop can be induced where data from individuals whose care has been influenced by the model is used to subsequently update the model for future individuals. Model-based treatment initiation may alter patient outcomes and therefore disrupt the relationship between the predictor variables and outcome itself, resulting in poorer model performance when the model is updated. [4,5] This is sometimes called performative prediction or the prediction paradox, and it has been suggested that QRISK3 is likely subject to this issue. [6,7]

Including any interventions in the model itself has been put forward as a way to mitigate the effects of performative prediction. [8] Previous work has shown that ignoring treatment in prognostic models leads to poor performance. [9] However, further research is needed to investigate treatment inclusion in the context of intervention feedback loops specifically and to provide quantitative evidence of the potential impact of this issue in practice. This study aims to use statistical simulation to investigate and quantify the potential impact of model-based treatment initiation on long-term model performance, as well as the further effect of including treatment in the model.

## Methods

Four scenarios were simulated, combining inclusion or exclusion of treatment as a predictor in the fitted model with presence or absence of an unmeasured confounding variable in the data-generating process.

For each individual, *i*, a binary adverse outcome *y*_*i*_ was drawn from a Bernoulli distribution using the probability determined by Eq. (1). These probabilities depended on a continuous predictor (*X*_*i*_) drawn from a standard normal distribution, binary treatment status (*T*_*i*_), and, where applicable, a binary unmeasured confounder (*U*_*i*_). The intercept was calculated empirically to achieve the specified outcome prevalence in an untreated population. The coefficient for the continuous predictor was fixed at 1, and the treatment coefficient was calculated empirically to achieve the specified average risk reduction. The confounder coefficient (*α*_3_) was fixed at 0.5.

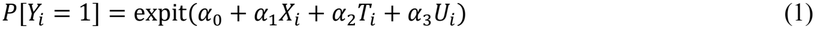

The parameters used for the simulation were motivated by CVD prevalence and intervention with statins. Accordingly, untreated outcome prevalence was set at 7%, predicted risk threshold for treatment was set at 10%, and risk reduction with treatment was set at 25%. [2,3,10]

Individuals were classified as high-risk if their predicted risk exceeded 10%. In scenarios without unmeasured confounding, high-risk individuals were always allocated treatment and low-risk individuals never received treatment.

In scenarios with unmeasured confounding, the confounder was randomly assigned to 20% of individuals and affected both outcome probability and treatment allocation. High-risk individuals with the confounder always received treatment; low-risk individuals with the confounder had a 50% chance of receiving treatment. High-risk individuals without the confounder had a 80% chance of receiving treatment; low-risk individuals without the confounder never received treatment.

For each scenario, the simulation proceeded as follows:

1. Data were generated for 10,000 individuals, including the continuous predictor, the unmeasured confounder where applicable, and potential outcomes under treatment and no treatment.
2. The sample is split 50:50. A model is fitted to the one half and internally validated with the other.
3. Data were generated for a new cohort of 10,000 individuals.
4. Prediction performance is calculated using the new *Y* values where treatment is not received.
5. Predicted risks were used to allocate treatment and update individual outcomes.
6. The model is refit using the latest data from these individuals only with their appropriate treatment statuses and matching outcomes.
7. Steps 3 to 6 are repeated for 10 cycles, with the updated model used in each successive cycle.
8. 200 repetitions of this process are carried out for each simulation scenario. Both prediction models were fitted using logistic regression. The first included only the continuous predictor *X*. The second included the continuous predictor and treatment status. Both omitted the unmeasured confounder.

Model performance was assessed by calibration-in-the-large (CITL), calibration slope, c-statistic, and proportion of individuals receiving a correct risk assignment based on their true risk relative to treatment threshold. The mean values of these figures are calculated in each cycle across the 200 repetitions.

Analyses were carried out in R version 4.5.2. [11] Code and expanded results are available in the following GitHub repository: https://github.com/SamPacynko/cpm-feedback-loop-sim

## Results

Figure 1 shows mean performance measures across simulation cycles. Cycle 0 represents validation of the initially developed model, and the first update occurs at the end of cycle 1. Performance measures from scenarios with and without unmeasured confounding are not directly comparable because the continuous predictor contributes differently to overall outcome variation. Nevertheless, changes across cycles and departures from ideal calibration remain informative.

**Figure 1.**
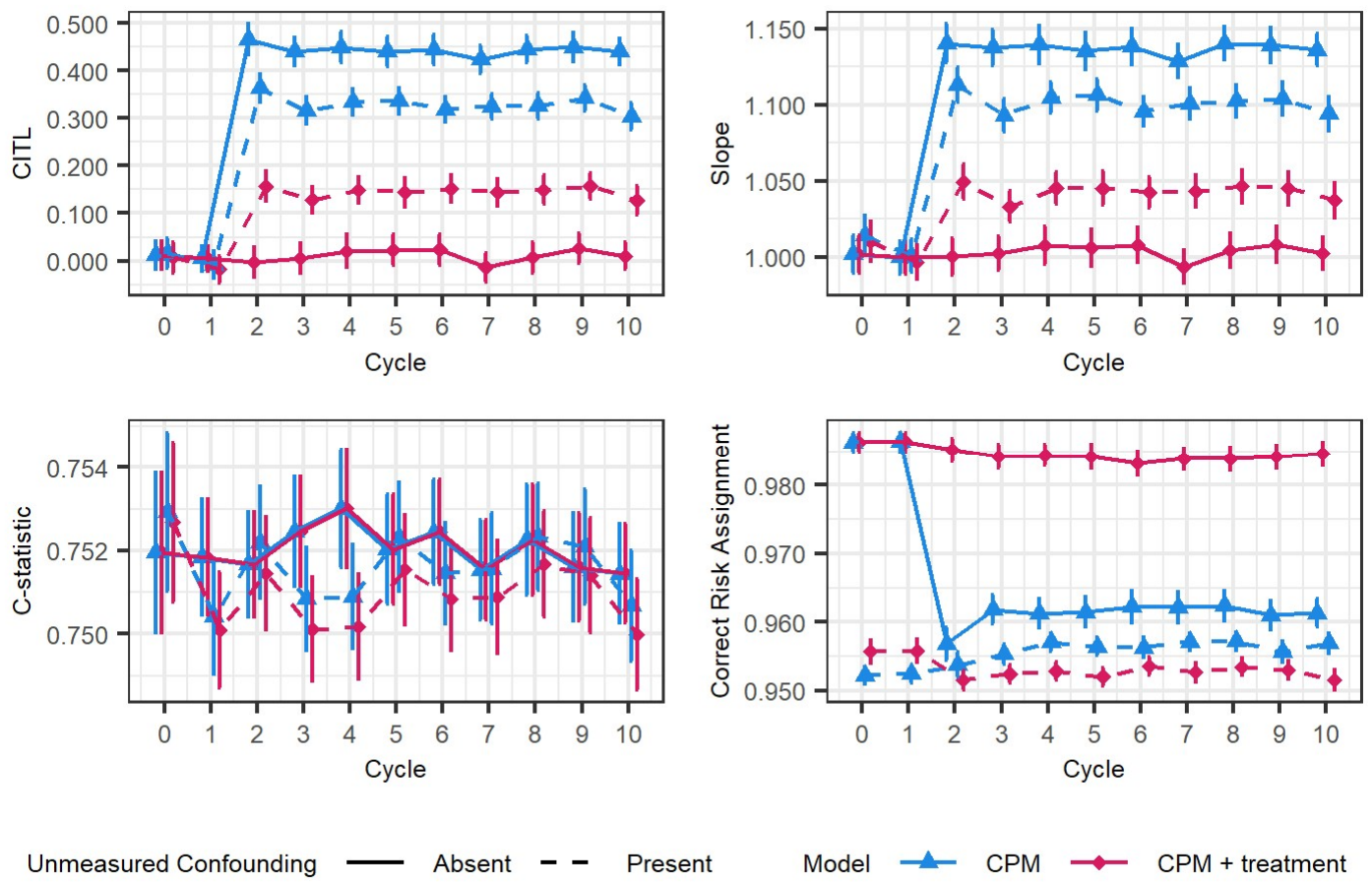
Average calibration-in-the-large (CITL), calibration slope, c-statistic, and proportion of individuals receiving a correct risk assignment with 95% confidence intervals at each update cycle for the two models of interest under presence/absence of unmeasured confounding.

In the absence of unmeasured confounding, updating the model without treatment as a predictor led to miscalibration. CITL indicated systematic underestimation of risk, while the calibration slope indicated that predictions were insufficiently extreme. Including treatment as a predictor maintained good calibration. In the presence of unmeasured confounding, both models exhibited miscalibration, irrespective of whether treatment was included.

Discrimination (c-statistic), remained largely unchanged following model updates for all four scenarios and little difference was observed between scenarios.

The proportion of individuals receiving a correct risk assignment was only retained at a high level when the model included treatment and there was no unmeasured confounding. Deterioration was observed post-model update when treatment was not included in the model and there was no unmeasured confounding. When there was unmeasured confounding, both models illustrated poorer risk assignment throughout.

## Discussion

Our findings support the importance of accounting for treatment when updating prediction models. Including treatment resulted in better performing model overall compared to when it is excluded. However, defining and recording treatment may be difficult in practice. Some interventions, such as dietary changes, may not be captured in routine data, while treatment may involve multiple interventions that vary according to patient circumstances and preferences.

Including treatment did not ensure good calibration in the presence of unmeasured confounding. Confounding is a known example of a causal blind spot in risk prediction.

[12] Given the complexity of real-world healthcare and the likelihood of all relevant factors being available for model development, it is probable that unmeasured confounding will always be an issue that needs to be considered in prediction modelling. Causal inference methods combined with risk prediction techniques may be required when confounding is suspected. [12]

The parameters used in this simulation are motivated by prediction for CVD outcomes and the results indicate that this setting has the potential to experience performance degradation when a similar feedback loop is present. In the scenario without unmeasured confounding, omitting treatment reduced the proportion of individuals receiving a correct risk assignment by approximately 2.5 percentage points. Given the prevalence of CVD, even a modest reduction in correct risk categorisation from a similar model could affect decisions for many individuals. [2] Further research is needed on post-implementation monitoring techniques for prediction models to allow identification of performance degradation due to feedback loops over time.

## Conclusions

When an implemented CPM is updated using data where treatment decisions have been affected by the model itself, the resulting feedback loop can result in decreased model performance. Including treatment in the model could help preserve these measures, but not completely when unmeasured confounding is present. The deterioration in performance observed across most scenarios can result in underestimation of risk and therefore missed opportunities for treatment. Causal inference methods may be needed to mitigate this issue further and more research is needed on model monitoring methods for identifying performance changes as they occur.

## Data Availability

All data produced are available online at https://github.com/SamPacynko/cpm-feedback-loop-sim

## Acknowledgements

Samantha Pacynko, Predoctoral Award NIHR500758, is funded by the NIHR for this research project. The views expressed in this publication are those of the authors and not necessarily those of the NIHR, NHS or the UK Department of Health and Social Care.

## Notes

### Competing Interest Statement

The authors have declared no competing interest.

